# COVID-19 (SARS-CoV-2) Ventilator Resource Management Using a Network Optimization Model and Predictive System Demand

**DOI:** 10.1101/2020.05.26.20113886

**Authors:** Samuel Billingham, Rebecca Widrick, Nathan J. Edwards, Sybil A. Klaus

**Affiliations:** The MITRE Corporation, Colorado Springs, CO 80910, USA; The MITRE Corporation, McLean, VA 22102, USA

**Keywords:** COVID-19, SARS, ventilators, medical device resources, pandemic management

## Abstract

The COVID-19 (SARS-CoV-2) pandemic is overwhelming global healthcare delivery systems due to the exponential spike in cases requiring specialty tests, facilities and equipment, including complex, precision devices like ventilators. In particular, the surge in critically ill patients has revealed a significant deficiency in regional availability of respiratory care ventilators. The authors offer a mathematical framework for ventilator distribution under scarcity conditions using an optimized network model and solver. The framework is interoperable with existing COVID-19 healthcare demand models and scales for different user-defined system sizes, including hospital networks, city, state, regional and national-scale prioritization. The authors’ approach improves current capabilities for medical device resource management within the existing incident command system while accounting for availability of devices, ventilation treatment time periods, disinfection and cleaning between patients, as well as shipping logistics time. The authors present a proof of concept using a high fidelity COVID-19 data set from Colorado, discusses how to scale nationally, and emphasizes the importance of applying ethical human-in-the-loop decision making when using this or similar approaches to managing medical device resources during epidemic emergencies.

## 1. Introduction

The global health challenges of combating the COVID-19 (SARS-CoV-2) pandemic have exposed constraints on healthcare systems including insufficient healthcare staffing, test kit availability, hospital ICU facilities, personal protective equipment, and availability of respiratory care devices such as ventilators. While traditional disaster incident command systems can account for broad resource management, the extreme ventilator resource demand and scarce supply during the pandemic have revealed shortcomings in current approaches to medical device resource management as described by Adelman [1] where intra-regional resource sharing must be considered as a risk mitigation. However the majority of mitigation efforts have been placed primarily on the emergency care of infected patients or on the strategy and tactics to reduce infection rates and not on the optimization of resource management for the complex demands within the U.S.

The complexity of medical device resource management during the COVID-19 pandemic provides a key opportunity for using computer based modeling and expert advisory systems to aid in the decision-making process of allocating resources as described by Beemer and Gregg [2]. In response to the pandemic in the United States, at least thirty predictive models on infection rates, ICU resource demand, and future ventilator demand have been developed. When these computational models are combined with real COVID-19 data, they allow for decisions to be made based on expected growth or demand. Additional models such as that presented by Polanco, et al. [3] aid in early detection of extreme demand on healthcare systems from severe respiratory disease epidemic outbreaks, however none of the models fully explore effective medical device resource management at regional or national levels.

This body of work presents a framework using a mathematical optimization for ventilator utilization and significantly aids in the management of scarce medical resources. And while the peak infection rates are gradually declining at the time of writing this manuscript, the approach will be highly applicable to any resurgence of COVID-19 or other future pandemics. The framework plans ventilator sharing within a region of interest by focusing on minimizing the number of patients not receiving a ventilator. The approach takes into account current ventilator inventories, addresses the arrival of new ventilators, typical ventilator treatment time periods, disinfection and cleaning time between patients, as well as shipping logistics time. The framework leverages existing COVID-19 infection prediction and healthcare demand models but can accommodate improved demand models or regional tailoring. The flexibility offered in the framework allows it to scale from local communities to counties, state, and nation. This work also presents a proof of concept using recent COVID-19 data from Colorado to demonstrate the algorithm along with a discussion on scaling to a national level. And finally the authors describe the importance of applying ethical decision making when using this or similar approaches to managing medical device resources during epidemic or pandemic emergencies.

## 2. COVID-19 Data Sets

Since the outbreak of the COVID-19 pandemic, a wide variety of data sets have been made available to the public. For example, the COVID-19 Dashboard by the Center for Systems Science and Engineering at Johns Hopkins University (JHU) provides case, recovery, and deaths counts globally by nation [4]. For the United States, this tool also breaks case counts down by county. Several states, counties, and municipalities within the United States are also providing COVID-19 data to the public, often with additional metrics.

Our research focuses on a proof-of-concept for ventilator sharing similar in nature to that described by Adelman and Gregg [1], but rather providing a scalable solution during peak and non-peak events. As such, we use the Colorado COVID-19 data provided by the Colorado Department of Public Health and Environment for this effort. Colorado has a population of nearly 6 million people and is made up of 64 counties. By using each county as a region for distributing ventilators, we are able to use a smaller set of data than would be required to distribute ventilators across the entire United States. Colorado’s data also provides many of the metrics needed to seed the model and demonstrate its capability. By using this smaller set of higher fidelity data, we are able to illustrate and provide proof of concept of our modeling construct, showing an optimal schedule of distributing ventilators across all Colorado counties in need. The modeling construct can then be scaled at regional, state or national levels. Sections 3 and 4 discuss all precise data needed to execute the model.

## 3. Modeling Ventilator Sharing

Mathematical optimization models are a representation of a set of choices to be made by a decision maker with the goal of maximizing or minimizing some objective. These decisions are subject to a set of constraints that express the limits on possible decision choices. For this work, time-phased decision variables are used in conjunction with time-phased balanced constraints over a fixed time horizon to allocate ventilators among facilities in order to meet demands in the entire region of interest. The following sections establish the notation and the structure of our model. The first three sections describe the sets, data parameters, and variables needed for the model with the final two sections establishing the objective and constraints for the model.

### 3.1. Sets

For our model we consider three sets. The first, 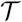 denotes time periods of interest when utilizing ventilators. The length of these time periods is inconsequential and could be set to whatever duration makes sense for the region. For our work we consider a time period to be one 24 hour period as most of the predictive models for ventilator demand predict daily values. The second, 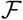 denotes the set of facilities in the region of interest utilizing ventilators. The last set, 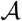 denotes the set of shipping routes, or arcs, between the facilities and is defined as the set of (*i, j*) pairs where 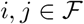 and *i* ≠ *j*.

### 3.2. Parameters

Parameters are the data items needed to execute the model. For our research we identified five parameters of interest. The parameter *P* denotes the intubation period for a patient and any ventilator reset time post extubation. While in practice these values will vary (i.e. intubation periods vary with patient age, gender, etc.), for a large scale planning model, it is helpful to assume away some of these sources of variability. Further research could explore the effects of variability in these parameters. The next two parameters *κ_i_* and *k_it_* are similar and denote the initial number of ventilators at facility *i* and any new ventilators arriving at facility *i* during time period *t* respectively. The first parameter *k* represents the ventilators that are native to the region while the second parameter *k_it_* represents ventilators not native to the region and are those supplied by increases in manufacturing, new purchases, arrivals from other regions, donations, etc. These new ventilators do not represent those shipped from another facility. We also assume that these new ventilators arriving in time period *t* will not be available for use until at least time period *t* + 1 as there will likely be some overhead time associated with in-processing the ventilator. The next parameter *d_it_* denotes the number of new patients at facility *i* needing ventilators in time period *t*. For our research, this parameter is based on predictive models and will be discussed later. The last parameter *s*_(*i, j*)_ is simply the shipping time, in time periods, on arc (*i, j*).

### 3.3. Variables

Variables represent the decisions that can be made when allocating ventilators. The first variable *y_it_* denotes the number of ventilators that are assigned to new patients at facility *i* during the time period *t*. The second variable *I_it_* represents the number of ventilators held in inventory at facility *i* at the end of time period *t*. The last variable *x_(i,j)_,_t_* denotes the number of ventilators that are shipped along arc (*i, j*) during time period *t*. This model assumes that the shipping logistics along the arc can be handled either by the facility or through a contracted shipper. Further research could explore the optimal shipping method although the pure logistics problem set is already well understood by large scale shipping companies such as FedEx or United Parcel Service (UPS).

### 3.4. Objective

The overall objective of our research is to minimize death of COVID-19 patients due to the lack of ventilators. To do this we make the assumption that if a patient does not receive a ventilator in the time period they need it, they will die of acute hypoxia or respiratory failure. To minimize death due to lack of ventilators we establish the following equation.

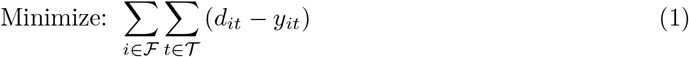

This objective function does not take into account any socio-political assumptions on the value of one life over another nor does it attempt to address disparity between facilities in the region. Future research could consider the effect of these assumptions on ventilator sharing.

For the modeling effort, a slightly different objective function is used to prevent excessive movement of ventilators in the schedule. That is, this objective as seen in Equation 2 simply penalizes the objective for moving the ventilator but not so much that the movement of ventilators will result in the loss of a life. This has the added benefit of creating a movement or transportation schedule that is relatively simple. For example, a 180-day planning period involving 28,775 cases spread across 64 counties in the state of Colorado produces a sharing schedule that involves only 117 shipments.

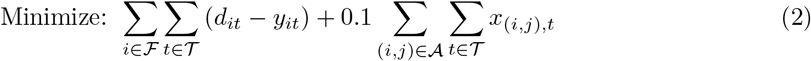

### 3.5. Constraints

All decisions are constrained including ventilator assignment and distribution following six constraints were identified.

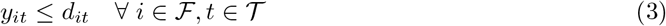

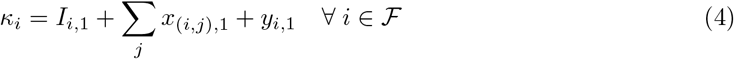

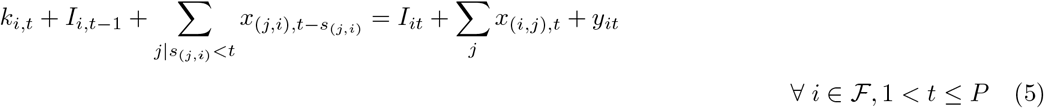

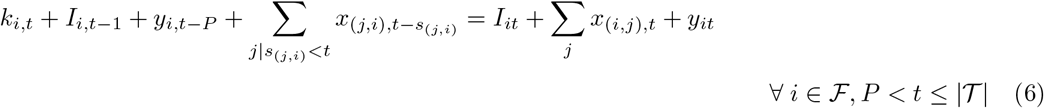

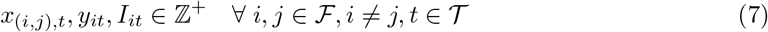

Constraint 3 ensures that ventilators assigned at a facility in a time period do not exceed the number of new patients arriving at the facility who need ventilator support during the time period. Constraints 4, 5, and 6 are the balance constraints and ensure that something is done with each available ventilator during the time period (assigned to patients, moved to a different facility, or held in inventory at the facility). These three constraints are needed as constraint 4 sets the initial conditions, constraint 5 holds during the time periods where extubations have yet to occur, and constraint 6 includes those time periods where extubations begin to occur. The last constraint 7 confines the decisions to non-negative integers.

## 4. Predicting Demand for Ventilators

The network optimization model is agnostic to the demand prediction model. Any model that can project new demand for ventilators can be used to populate the demand signal in the network optimization. The best available forecasting model for a specific region should be used to project demand.

To demonstrate our modeling construct, we model the demand projections, *d_it_*, using a standard Susceptible, Infected, Recovered (SIR) model to project new hospital admissions due to COVID-19 with a fixed proportion of these admissions requiring ventilators. The standard SIR model derived from Kermack-McKendrick [5] is defined by the following set of equations and constants where *t* represents the time period of interest.

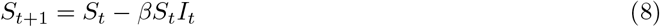

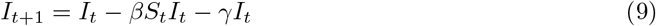

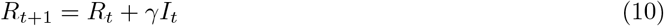

where

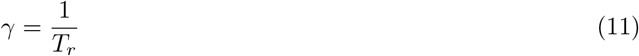

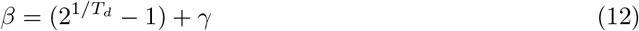

The constants, *T_d_* and *T_r_* represent doubling time and recovery time respectively and also aid in calculating the basic reproduction number (*R*_0_) of a disease. *R*_0_ represents the expected number of cases generated by a single case of a disease and indicates how contagious an infections disease is. Higher values of *R*_0_ indicate more rapid growth with *R*_0_ ≥ 1 indicating that the disease will spread. For a notional disease, a doubling time of 4 days (*T_d_* = 4) and recovery time of 14 days (*T_r_* = 14) implies

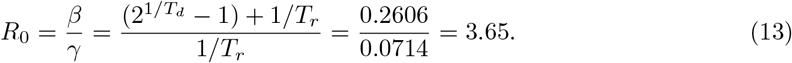

This means each infected person, on average, will infect 3.65 other people. Applications of the SIR model can also be adjusted to account for social distancing measures by making adjustments to the reproduction number over time, i.e. *R_t_*. For our notional disease, if we assume social distancing measures reduce population interaction by 30% beginning on day *t*, this will increase the disease’s doubling time to 6.6 days, resulting in *R_t_* = 2.55.

For our research we use the COVID-19 Hospital Impact Model for Epidemics (CHIME) developed by the Penn Medicine Predictive Healthcare team [6]. CHIME implements a SIR model that allows hospitals to enter information about the population in their entrapment area and modify assumptions around the spread and behavior of COVID-19. CHIME applies to a population, specifically a hospital’s entrapment area, it is expandable to any population. For example, Figure 1 shows how CHIME can be applied at the county level and displays new ventilator demand in the 10 most infected counties in the State of Colorado. This figure assumes a 30% reduction in social contact, one ventilator needed per hospitalization, and *R*_0_ = 2.55. Figure 1 also demonstrates how demand for ventilators will peak at different times during the COVID-19 pandemic. This further supports the thesis that if ventilators are shared between populations within a region of interest more patients will receive needed ventilators.

**Figure 1:**
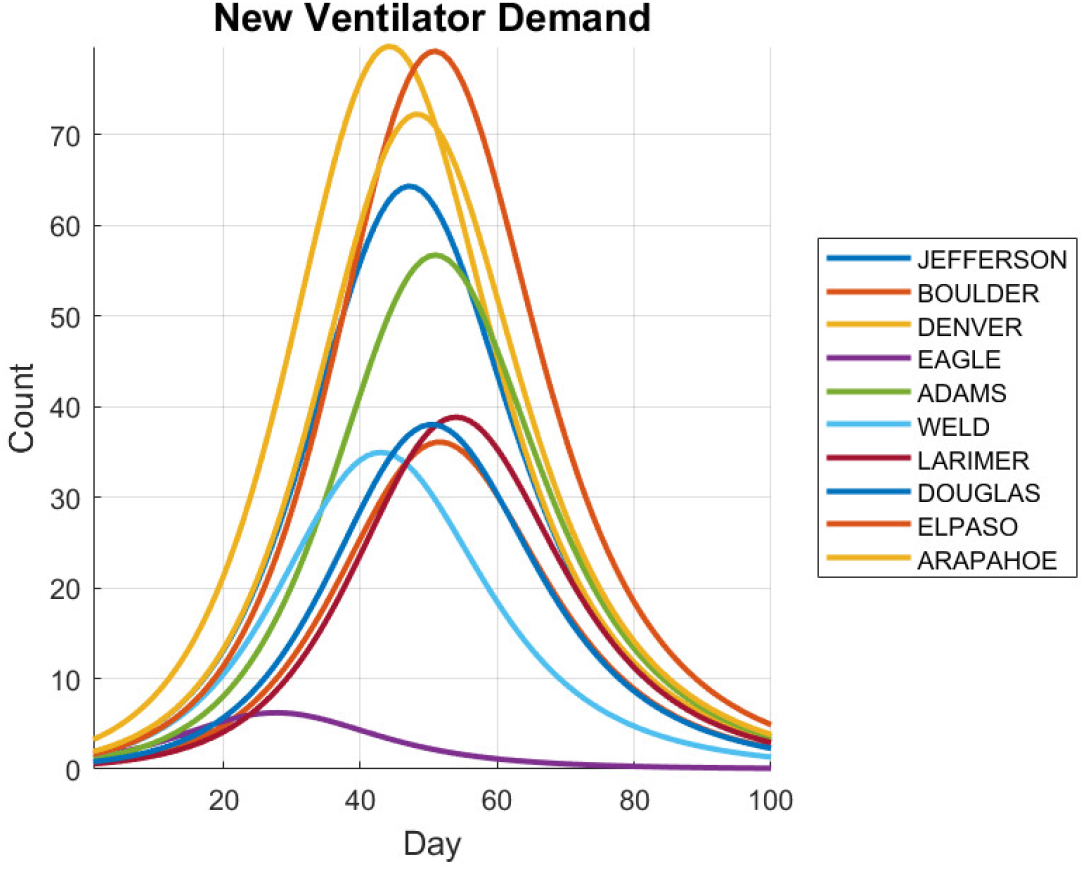
Predicted daily ventilator demand for Colorado counties with highest COVID-19 infection counts on March 31, 2020. Assumes 30% population interaction reduction due to social distancing measures.

CHIME also allows for sensitivity analysis by changing the underlying assumptions required to predict new ventilator demand. Figure 2 shows the expected ventilator demand in Denver County, Colorado throughout the COVID-19 pandemic adjusted for differing reductions in social contact. For example, if planners assume that social distancing measures result in a 30% reduction in population interaction and social contact in Denver County, then the model will surge ventilators to the county in the early days of the pandemic. However, if the measures had instead resulted in a 50% reduction in population interaction the ventilator surge will arrive early and be unavailable in other counties. Since the social distancing parameters can be adjusted and modeled quickly, the impact of varying assumptions can be compared and better sharing strategies developed. One potential limitation of the CHIME model is that it is not calibrated at the local level.

**Figure 2:**
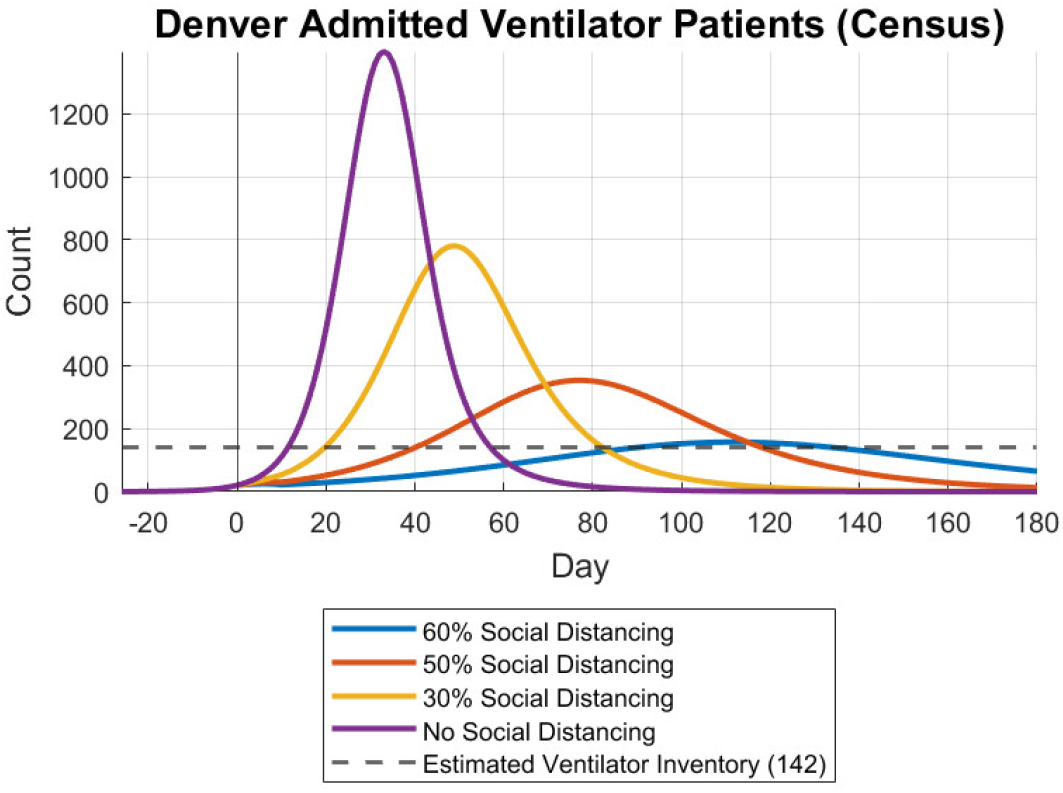
Effect of social distancing assumptions on predicted daily ventilator demand for Denver County on March 31, 2020.

Institute for Health Metrics and Evaluation (IHME) COVID-19 Projections model [7] is another high fidelity model that takes into account when social distancing policies go into effect at the state and global levels, which has a strong impact on the infections over time. IHME’s model could also be considered for predicting ventilator demand over time.

## 5. Demonstration

This section demonstrates how the prediction and ventilator sharing models work together to minimize patient death due to lack of ventilators. To demonstrate, we utilize population data, case counts, and ventilator inventory estimates from each county in the State of Colorado with a confirmed COVID-19 infection as of March 31, 2020. The population estimates, case counts, and hospitalization estimates are used to predict future demand for ventilators. The population estimate is also used to estimate the number of ventilators in each county with a ratio of 19.7 ventilators per 100,000 people [8]. These ventilator counts serve as the *κ_i_* values in the ventilator sharing model.

The counties serve as proxies for facilities as the data can be readily accessed and can aid in planning at a regional level. Future research could isolate the planning to individual hospitals. Table 1 shows the estimated populations, case counts, estimated hospitalizations, and estimated ventilator inventory from each of the Colorado counties with at least one estimated COVID-19 hospitalization.

**Table 1:** Population, case counts, estimated hospitalizations, and ventilator inventory estimates from Colorado counties with at least one estimated COVID-19 hospitalization on March 31, 2020.

| County | Pop. | Cases | Hospitalized | Vent. Inv. |
| --- | --- | --- | --- | --- |
| Denver | 717797 | 539 | 97 | 142 |
| El Paso | 714395 | 286 | 51 | 141 |
| Arapahoe | 651342 | 333 | 60 | 129 |
| Jefferson | 579491 | 304 | 55 | 115 |
| Adams | 511473 | 181 | 32 | 101 |
| Larimer | 350362 | 99 | 18 | 70 |
| Douglas | 342842 | 141 | 25 | 68 |
| Boulder | 325476 | 107 | 19 | 65 |
| Weld | 314251 | 255 | 46 | 62 |
| Pueblo | 167116 | 21 | 4 | 33 |
| Mesa | 153630 | 14 | 3 | 31 |
| Broomfield | 69453 | 20 | 4 | 14 |
| Garfield | 59807 | 33 | 6 | 12 |
| La Plata | 56403 | 23 | 4 | 12 |
| Eagle | 54863 | 227 | 41 | 11 |
| Montrose | 42260 | 13 | 2 | 9 |
| Summit | 30973 | 20 | 4 | 7 |
| Morgan | 28503 | 4 | 1 | 6 |
| Elbert | 26218 | 5 | 1 | 6 |
| Routt | 25683 | 17 | 3 | 6 |
| Teller | 25060 | 7 | 1 | 5 |
| Logan | 21854 | 6 | 1 | 5 |
| Chaffee | 20028 | 17 | 3 | 4 |
| Park | 18557 | 3 | 1 | 4 |
| Otero | 18364 | 3 | 1 | 4 |
| Pitkin | 17879 | 30 | 5 | 4 |
| Gunnison | 17173 | 82 | 15 | 4 |
| Grand | 15474 | 4 | 1 | 4 |
| Moffat | 13181 | 4 | 1 | 3 |
| Rio Grande | 11226 | 5 | 1 | 3 |
| Clear Creek | 9663 | 4 | 1 | 2 |
| San Miguel | 8177 | 4 | 1 | 2 |
| Costilla | 3809 | 3 | 1 | 1 |
| Baca | 3547 | 3 | 1 | 1 |

Populations, positive COVID-19 case counts, and total Colorado COVID-19 hospitalizations are provided by the Colorado Department of Public Health and Environment. Hospitalizations by county are estimated using the total hospitalizations and the proportion of positive cases in each county.

To begin the demonstration we adapt CHIME in MATLAB to batch process demand predictions for all 34 counties in Colorado with at least one estimated hospitalization due to COVID-19. The data of interest are new daily ventilator admissions. Table 2 shows the model inputs and assumptions used to predict ventilator demand. Settings were kept the same as the default settings in CHIME with the exception of regional population and currently hospitalized, which were varied based on the regions of interest. The settings can be easily modified as new data emerges regarding COVID-19.

**Table 2:** CHIME inputs & settings

| Variable | Setting |
| --- | --- |
| Regional Population | see Table 1 |
| Hospital Market Share | 100% |
| Currently Hospitalized COVID-19 Cases | see Table 1 |
| Doubling Time (Days) | 4 |
| % Reduction in Population Interaction (Social Distancing) | 30% |
| Hospitalizations (% Infected) | 2.5% |
| ICU (% Infected) | 0.75% |
| Ventilated (% Infected) | 0.5% |
| Infectious Days | 14 |
| Average Days Hospitalized | 7 |
| Average Days in ICU | 9 |
| Average Days on Ventilator | 10 |
| Number of Days to Project | 180 |

Figure 3 demonstrates the new demand predictions generated for Denver County. Day 0 in the figure represents March 31, 2020. For our model we assume social distancing measures went into effect on this date and accounts for the slight dip in new hospital admissions early on. In order to adequately account for partial values in the prediction we round all partial values up to the nearest integer (e.g., 2.02 new admissions → 3 new admissions). And while this is a slight over-prediction, the authors believe that this provides the best means to conservatively handle these types of values especially since fractional patients cannot be a real value.

**Figure 3:**
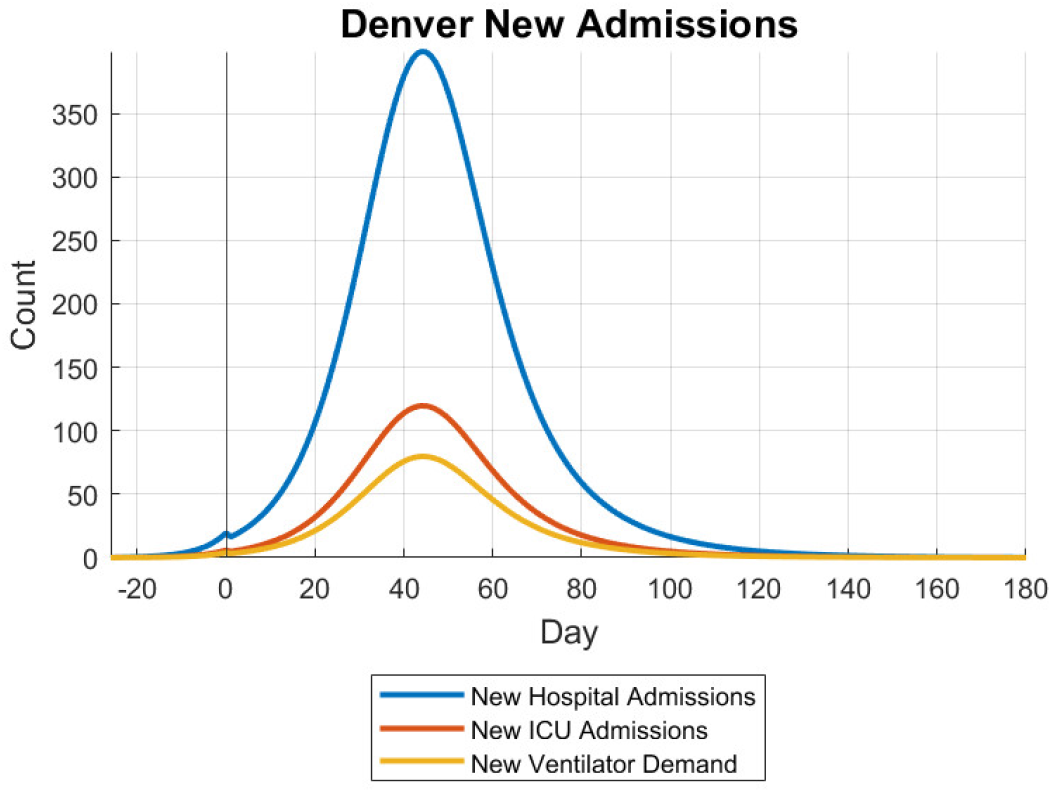
Denver 30% Social Distancing New Admissions - March 31, 2020

Once generated, the predicted ventilator demands provide the *d_it_* values needed for the ventilator distribution model. Note that two parameters needed for the CHIME model are also needed for the ventilator sharing model. First the average number of days on a ventilator equates to the ventilator intubation and reset period. We use *P* = 10 in this ventilator distribution model demonstration, although *P* can be adjusted to a value that is consistent with current clinical data on ventilator treatment periods. Second, the number of days to project equates to the number of time periods to plan, in our case 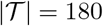. This leaves two remaining parameters to discuss, shipping times and new ventilators.

For shipping times in our demonstration we simply use the geographic coordinates for the center of each county and measure the straight-line distance between all counties, divide by a estimated travel distance per day and round up. Rounding up occurs to force at least one day for shipping. This assumption is based on our belief that the ventilator, if shipped, will take at least a day to pull out of service at the facility, go through processing to clean and inventory, be packaged for shipping, shipped, and then processed on the receiving end before being put back into service. Further work is needed to precisely calculate shipping times and facilities may have to work with their local shipping vendors to establish these times.

Lastly, new ventilators being introduced into the system are accounted for. Since this team does not have accurate counts nor expected arrival dates for any new ventilators in Colorado (either purchases or national stockpile), we provide new ventilator counts for demonstration purposes only. For the demonstration the team will use the following values; 100, 250, and 500 new ventilators arriving in Denver (Denver is the main transportation hub for Colorado) on the 6^th^, 13^th^, and 20^th^ of April respectively.

Now that the parameters have been defined we demonstrate the sharing model utilizing two different scenarios. The first scenario shows the predicted death rate due to lack of ventilators under three conditions: (1) no sharing, (2) sharing, and (3) sharing with new ventilators. The second demonstrates how sensitivity analysis can be conducted as the social distancing parameters are adjusted to show how ventilator sharing is robust to changes in assumptions.

For the first scenario the sharing model is run under three conditions. The first condition, no sharing, indicates that there will be 17,678 deaths due to the lack of ventilators. The second condition, sharing, indicates that there will be no deaths due to lack of ventilators. To accomplish this 117 ventilator shipping actions will occur throughout the 180 day planning cycle. The last scenario also results in no deaths due to lack of ventilators as simply injecting more ventilators into a system that already contains enough to meet demand throughout the planning horizon produces the same result. However, the additional ventilators do produce a sharing plan that only involves 62 shipping actions. Table 3 provides a snapshot of the shipping schedule for condition 2.

**Table 3:**
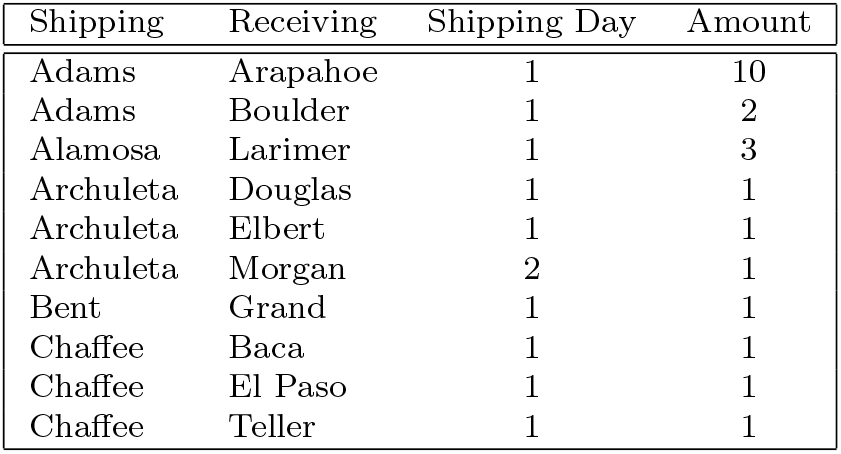
First ten entries of the ventilator sharing schedule from scenario 1 condition 2.

| Shipping | Receiving | Shipping Day | Amount |
| --- | --- | --- | --- |
| Adams | Arapahoe | 1 | 10 |
| Adams | Boulder | 1 | 2 |
| Alamosa | Larimer | 1 | 3 |
| Archuleta | Douglas | 1 | 1 |
| Archuleta | Elbert | 1 | 1 |
| Archuleta | Morgan | 2 | 1 |
| Bent | Grand | 1 | 1 |
| Chaffee | Baca | 1 | 1 |
| Chaffee | El Paso | 1 | 1 |
| Chaffee | Teller | 1 | 1 |

For the second scenario we demonstrate a sensitivity analysis using the model setup parameters as presented except we will vary the social distancing (or reduction in social contact) parameter when predicting new ventilator demands. This demonstration shows how planning assumptions can be varied allowing planners to understand the associated risks and to adjust their plans as these assumptions change. The first condition for this demonstration will serve as the baseline and use the 30% reduction in social contact parameter, as well as sharing with no new ventilators. The second condition will increase the social distancing to 50%, highlighting the movement of peak demand further into the planning horizon. Lastly, the social distancing parameter will begin with 30%, increase to 50% on day 15 of the planning horizon, then decrease back to 30% on day 61 of the planning horizon. This attempts to emulate an initial social distancing period, followed by a period of tightening restrictions, and a return to looser restrictions. This demonstrates the robustness of the method under changing and dynamic assumptions. Each run condition will assume the arrival of new ventilators into the region.

The first condition of the second scenario assumes a 30% reduction in social contact and yields zero deaths due to not having a ventilator and produces 62 shipping actions. The second scenario assumes a 50% reduction in social contact and also yields zero deaths due to not having a ventilator. The difference is in the sharing of ventilators and yields a sharing schedule that involves 64 shipping actions. The final scenario varies the social distancing parameter over time and again yields zero deaths due to lack of ventilators. These scenarios show that the initial planning assumptions can be modified throughout the course of the pandemic and schedules adjusted. Also while the social distancing assumptions were modified in the scenarios, sensitivity analysis is not limited with this parameter. In fact, any of the parameters used in the sharing model or the prediction model can be varied and their impacts explored.

## 6. Scaling Considerations

While our mathematical framework and approach can accommodate improved ventilator demand models and other constraints, it is important to recognize the various regional aspects for scaling (up or down) to specific problem sets. The presented proof of concept is based on publicly available COVID-19 data for the state of Colorado that serves as a general proxy for both ventilator demand and shipping in other regions, but in order to make healthcare resource allocation decisions for local communities, counties, state and national levels there are a number of tailored inputs that should be considered. For example, each region is likely to have different assumptions of ventilator prioritization and maintenance cycles within single hospital, city, county, state, etc. Additionally the presented model generally optimizes for nearest neighbor (non-mathematical competitors), but might require human-in-loop decisions based on existing healthcare partnerships or other sources of medical devices or disposable ventilator service items.

One of the key elements of this framework that lends itself to scalability is the flexibility of parameter *s*_(*i, j*)_, the shipping time along transportation route or arc *(i,j*). The shipping time needs to account for intra-regional and inter-regional transportation, roadways, airways, railways, waterways, and also logistics midway nodes or transportation hubs. While our proof of concept uses GPS lat/long of mid-points for each county, the details of shipping time and logistics are best provided by the available commercial companies such as FedEx, UPS, DHL, or other express logistic services who offer an application programming interface (API). In some cases the transportation estimation may be provided by a local staff or public health employee who will be individually transporting the medical devices following an optimized route provided by a GPS traffic mapping application.

Additional considerations for scaling this approach to a national level are the computational needs, both hardware and software, for computing the distribution solution of a complex national system. If computer calculations are too slow, then solutions will not account for current event data or may be obsolete by the time the output is available. For the ventilator sharing model, scenarios were run on a Sun Fire x4150 with 2 Intel Zeon E5440 2.83GHz 8 core processors with 16 GB RAM. CPLEX 12.10.0.0 served as the solver and AMPL was used to setup and implement the sharing model into CPLEX. All scenarios presented solve to optimality within 5 minutes.

For the demand prediction model, projections were done on a Dell Latitude 7480 Intel Core i7-7600U CPU @ 2.8GHz, 2901Mhz, 2 Cores, 4 Logical Processors and 16GB RAM. For Colorado, 34 out of the 64 counties have at least one estimated hospitalization as of March 31, 2020. Predicting demand curves for all 34 counties over 180 days using MATLAB R2019a takes 1.12 seconds. To predict all 64 counties over 180 days takes 1.15 seconds. However these time frames are very dependent on the hardware used and the scale of the sharing model (64 counties and 180 days to plan).

There is no guarantee that the optimal solution for larger regions or the national-level will solve within this documented time frame on the described hardware. It is suggested to consider the use of high performance computing systems that might be available through government or local university resources for larger problem sets desiring a higher fidelity solution within hospital networks, counties, or regions with more cities.

## 7. Ethical Considerations of Using a Scarce Resource Allocation Framework

During a catastrophic public health crisis, a scarce resource allocation framework can aid health officials more effectively use medical resources to do the greatest good for the greatest number of patients [9, 10, 11]. However, healthcare decisions based solely on computational predictive models will be limited by their assumptions, and may not be grounded in standard healthcare practices nor would they account for ethical decisions that take into account situational considerations. Fortunately there are many different guidelines regarding the ethical criteria to inform the creation of an allocation framework for ventilators or critical care resources [12, 13]. The goals of an allocation framework often include providing meaningful access and individualized assessments based on the best available medical evidence for all patients. In addition, most decision makers agree a successful allocation effort requires public trust and cooperation through transparent and inclusive participation in the process.

The use of an allocation framework generated with community engagement prior to a crisis can help ensure no patient is denied care based on stereotypes, assessments of quality of life or judgments about their ‘worth’ based on the presence or absence of disabilities or other factors. There is significant caution to consider not using categorical exclusion criteria because they are often too rigid during a dynamic crisis and can be associated with discrimination. Other considerations include: periodic reassessments of patients to maximize population health outcomes; prioritization of individuals with vital functions during a pandemic (e.g. essential workers and healthcare workers). Potential methods to generate an allocation of resources are outlined by White et. al [12, 11]; we adapt those to the use of our mathematical framework for ventilator distribution:

1. Create triage teams to ensure consistent decision making on the availability and needs of scarce critical care resources at the local healthcare facility;
2. Establish criteria for initial allocation of incoming ventilators; and
3. Establish reassessment criteria to determine whether ongoing provision of ventilators are justified for individual patients, and to ensure the extensive regional demand for ventilator resources are not biasing individual patient healthcare decisions.

Another key consideration is that any implemented solution based on the method described in the mathematical framework should use demand values based on actual healthcare data or in combination with predictive models that fully describe their assumptions. Some states like Colorado and other regions offer this data, while others may not. It is important to consider the fusion of real and predictive data to guide ethical decision making but not to provide definitive healthcare resource management.

## 8. Conclusions

Our work demonstrates a mathematical framework for ventilator distribution under scarcity conditions using an optimized network model and solver and shows when to transport ventilators and to which locations while accounting for availability of devices, ventilation treatment time periods, disinfection and cleaning between patients, as well as shipping logistics time. While the proof of concept used Colorado data with GPS center point locations and a generalized ventilator predicative demand model, it represents elements of all healthcare systems. Our work also discusses some of the scaling considerations that require regional or situational adjustments for actual geographic locations, shipping logistics and a daily or 12-hour updates to the ventilator demand signal with real data. We also emphasize the importance of applying ethical human-in-the-loop decision making when using this or similar computational predictive model approaches to managing medical device resources during epidemic emergencies. The foundations of this work can also apply to other scarce medical resource challenges. Future work should investigate the nuances of applying this approach to special hospital health networks in addition to leveraging synthetic patient healthcare data [14] as a more accurate predictor of emerging resource needs.

## Data Availability

The datasets generated for this study are available to U.S. national, state and regional agencies as well as healthcare systems on request to the corresponding author.

## Author Contributions

Mr. Billingham developed the network optimization model and proof of concept. Ms. Widrick developed the Ventilator demand model dataset for Colorado, Mr. Edwards contributed to the use-case and background research. Dr. Klaus provided key discussion on the ethics and use of computer-based models in a healthcare setting.

## Acknowledgments

We would like to acknowledge CHIME at Penn Medicine for making their model available for public use. We could not have reproduced their logic and generated the demand curves seen in this paper in such a rapid turnaround without their methodology and downloadable source code.

## References

1. Adelman D. Thousands Of Lives Could Be Saved In The US During The COVID-19 Pandemic If States Exchanged Ventilators. Health Affairs 2020;:10.1377/hlthaff.2020.00505URL: https://www.healthaffairs.org/doi/full/10.1377/hlthaff.2020.00505. doi:10.1377/hlthaff.2020.00505; publisher: Health Affairs.

2. Beemer BA, Gregg DG. Advisory Systems to Support Decision Making. In: Burstein F. W. Holsapple C, eds. Handbook on Decision Support Systems 1: Basic Themes. International Handbooks Information System; Berlin, Heidelberg: Springer. ISBN 978–3-540-48713-5; 2008:511-27. URL: https://doi.org/10.1007/978-3-540-48713-5_24. doi:10.1007/978-3-540-48713-5_24.

3. Polanco C, Castañón-González JA, Macias AE, Samaniego JL, Buhse T, Villanueva-Martínez S. Detection of Severe Respiratory Disease Epidemic Outbreaks by CUSUM-Based Overcrowd-Severe-Respiratory-Disease-Index Model. Computational and Mathematical Methods in Medicine 2013;2013. URL: https://www.ncbi.nlm.nih.gov/pmc/articles/PMC3771461/. doi:10.1155/2013/213206.

4. Center for Systems Science and Engineering (CSSE) at Johns Hopkins University (JHU). COVID-19 Dashboard. https://coronavirus.jhu.edu/map.html; 2020. URL: https://coronavirus.jhu.edu/map.html.

5. Anderson RM, May RM. Population biology of infectious diseases: Part I. Nature 1979;280(5721):361–7. URL: https://www.nature.com/articles/280361a0. doi:10.1038/280361a0; number: 5721 Publisher: Nature Publishing Group.

6. Predictive Healthcare at Penn Medicine. COVID-19 Hospital Impact Model for Epidemics (CHIME). https://penn-chime.phl.io/; 2020. URL: https://penn-chime.phl.io/.

7. IHME COVID-19 Projections. https://covid19.healthdata.org/; 2020. URL: https://covid19.healthdata.org/; library Catalog: covid19.healthdata.org.

8. Rubinson L, Vaughn F, Nelson S, Giordano S, Kallstrom T, Buckley T, Burney T, Hupert N, Mutter R, Handrigan M, et al. Mechanical ventilators in us acute care hospitals. Disaster Medicine and Public Health Preparedness 2010;4(3):199–206. doi:10.1001/dmp.2010.18.

9. Childress JF, Faden RR, Gaare RD, Gostin LO, Kahn J, Bonnie RJ, Kass NE, Mastroianni AC, Moreno JD, Nieburg P. Public health ethics: mapping the terrain. The Journal of Law, Medicine & Ethics: A Journal of the American Society of Law, Medicine & Ethics 2002;30(2):170–8. doi:10.1111/j.1748-720x.2002.tb00384.x.

10. Gostin L. Public Health Strategies for Pandemic Influenza: Ethics and the Law. JAMA 2006;295(14):1700–4. URL: https://jamanetwork.com/journals/jama/fullarticle/202648. doi:10.1001/jama.295.14.1700; publisher: American Medical Association.

11. White DB. A Model Hospital Policy for Allocating Scarce Critical Care Resources 2020;URL: https://ccm.pitt.edu/?q=content/model-hospital-policy-allocating-scarce-critical-care-resources-available-online-now.

12. White DB, Lo B. A Framework for Rationing Ventilators and Critical Care Beds During the COVID-19 Pandemic. JAMA 2020;URL: https://jamanetwork.com/journals/jama/fullarticle/2763953. doi:10.1001/jama.2020.5046.

13. Biddison ELD, Gwon HS, Schoch-Spana M, Regenberg AC, Juliano C, Faden RR, Toner ES. Scarce Resource Allocation During Disasters: A Mixed-Method Community Engagement Study. CHEST 2018;153(1):187–95. URL: https://journal.chestnet.org/article/S0012-3692(17)31393-4/abstract. doi:10.1016/j.chest.2017.08.001; publisher: Elsevier.

14. The MITRE Corporation. Synthea™ Synthetic Patient Generator. https://synthetichealth.github.io/synthea/; 2020. URL: https://synthetichealth.github.io/synthea/.

